# The ansa subthalamica: a neglected fiber tract

**DOI:** 10.1101/19002352

**Authors:** Eduardo Joaquim Lopes Alho, Ana Tereza Di Lorenzo Alho, Andreas Horn, Maria da Graca M. Martin, Brian L Edlow, Bruce Fischl, József Nagy, Erich T Fonoff, Clement Hamani, Helmut Heinsen

**Author notes:** Corresponding Author Dr. Eduardo Joaquim Lopes Alho, MD, PhD, Rua Pamplona, 1585, apto 53, cep 01405-002 São Paulo-SP Brazil.

## Abstract

**Background:** The pallidofugal pathways are classically subdivided into ansa lenticularis, lenticular fasciculus, and subthalamic fasciculus. In addition to these three subsystems, we characterize an anatomical structure that connects the antero-medial pole of the subthalamic nucleus to the ventral portions of the pallidum, both related to limbic processing of information. This bundle has been previously considered to form a part of the ansa lenticularis, however, it shows striking differences on histology and MRI features compared to the ansa lenticularis, and therefore we suggest to denominate it *ansa subthalamica*.

**Objectives:** To describe the ansa subthalamica as a different structure than the ansa lenticularis, that can be recognized by different methods (histology, high-field MRI and connectome tractography), including current 3T clinical imaging.

**Methods:** A complete human brain was histologically processed and submitted to registration procedures to correct for tissue deformations and normalization to MNI space. Coordinates of histological structures were then comparable to high-field (7T) *post-mortem* and *in vivo* MRIs, 13 pre-operative 3T imaging of parkinsonian patients and normative connectome tractography. Mean intensity gray values for different structures were measured in Susceptibility-Weighted Images.

**Results:** It was possible to characterize this structure with different methods and there was significant difference in signal intensity in the ansa subthalamica (hypointense), compared to the ansa lenticularis (hyperintense).

**Conclusions:** The ansa subhtalamica may represent the anatomical pathway that connects limbic regions of the STN and pallidum, and should be investigated as a possible substrate for limbic effects of stereotactic surgery of the subthalamic region.

## Introduction

The first detailed description of the pallidofugal pathways dates from 1895 when Constantin von Monakow made observations based on histological sections through diseased human specimens. He subdivided the major fiber systems of the basal ganglia into ansa lenticularis (*Linsenkernschlinge* or lenticular nucleus loop), the lenticular fasciculus, and the subthalamic fasciculus. Subsequent animal research with anterograde degeneration methods and axonal tracing demonstrated that these three subsystems arise from different pallidal regions, although this concept has recently been questioned^1^. The AL originates in the lateral division of the globus pallidus internus, with a rostral and medial trajectory, curving around the internal capsule in a medial, posterior, and superior course to reach the H field of Forel and then the thalamus^2^. Fibers from LF originate in the medial subdivision of the GPi and traverse dorsomedially, perforating the internal capsule, and thus forming the anatomical structure known as Edinger’s comb system^3^. The SF originates mainly from the GPe, implementing the motor circuit’s “indirect pathway” as it was described by Alexander in 1990^4^.

According to the tripartite functional subdivision of the STN, the ventromedial aspect of the nucleus processes mainly limbic information ^5^. Neurons of the medial tip of the STN project to the ventral pallidum in non-human primates and cats^6,7^, however these connections are poorly demonstrated in humans.

*Post-mortem* techniques for white matter visualization include anatomical dissection of frozen brains (Klinger’s method), histological myelin staining, polarized light imaging, polarization-sensitive optical coherence tomography^8^ and structural tensor analysis ^9^. Diffusion tensor imaging (DTI) and tractography, introduced in the early 1990s^10,11^, allows an *in-vivo* connectivity mapping of the human brain ^12^. More recently, our group has studied serial high thickness histological (430µm) slices under light- and dark-field microscopy. By increasing the contrast between fiber bundles and nuclear boundaries^13^, this method facilitates the delineation and documentation of neural structures in human *post-mortem* tissue. *In-situ* MRI allowed registration of the whole histological brain to its own scan before the occurrence of tissue processing deformations, and normalization to MNI space. As such, this high-resolution and high-contrast histological atlas could be directly used in both populational studies^14^ or to map individual patient’s anatomy^15–17^. This methodology improves the visualization and assists volumetric reconstruction of complex anatomical structures and fiber tracts.

With the advance of neuromodulation and DBS techniques targeting neuropsychiatric disorders, it is important to understand this intricate anatomical region in humans and envision potential therapeutic strategies. Great therapeutic benefits of DBS were recorded in the last decades for various movement disorders, with significant improvement in quality of life of thousands of patients worldwide. This fact motivated countless scientific studies involving the subthalamic nucleus and the surrounding areas, as one of the most studied regions in the brain with important discoveries and clinical correlations. In addition to the three subsytems of fibers described above, we characterize an anatomical structure that connects the antero-medial pole of the STN to the ventral portions of the pallidum. It differs in topography, histological features and magnetic properties from the classically described pallido-subthalamic pathways (Fig. 1). The bundle has been previously considered to form a subset or part of the ansa lenticularis^18^. However, it shows striking differences on histology and MRI compared to this bundle. The material presented here suggests that it should receive nomenclature that differentiates it from the ansa lenticularis (which largely innervates the thalamus). Since it terminates in the subthalamic nucleus, we suggest calling this bundle *ansa subthalamica* or subthalamic ansa.

**Fig.1.**
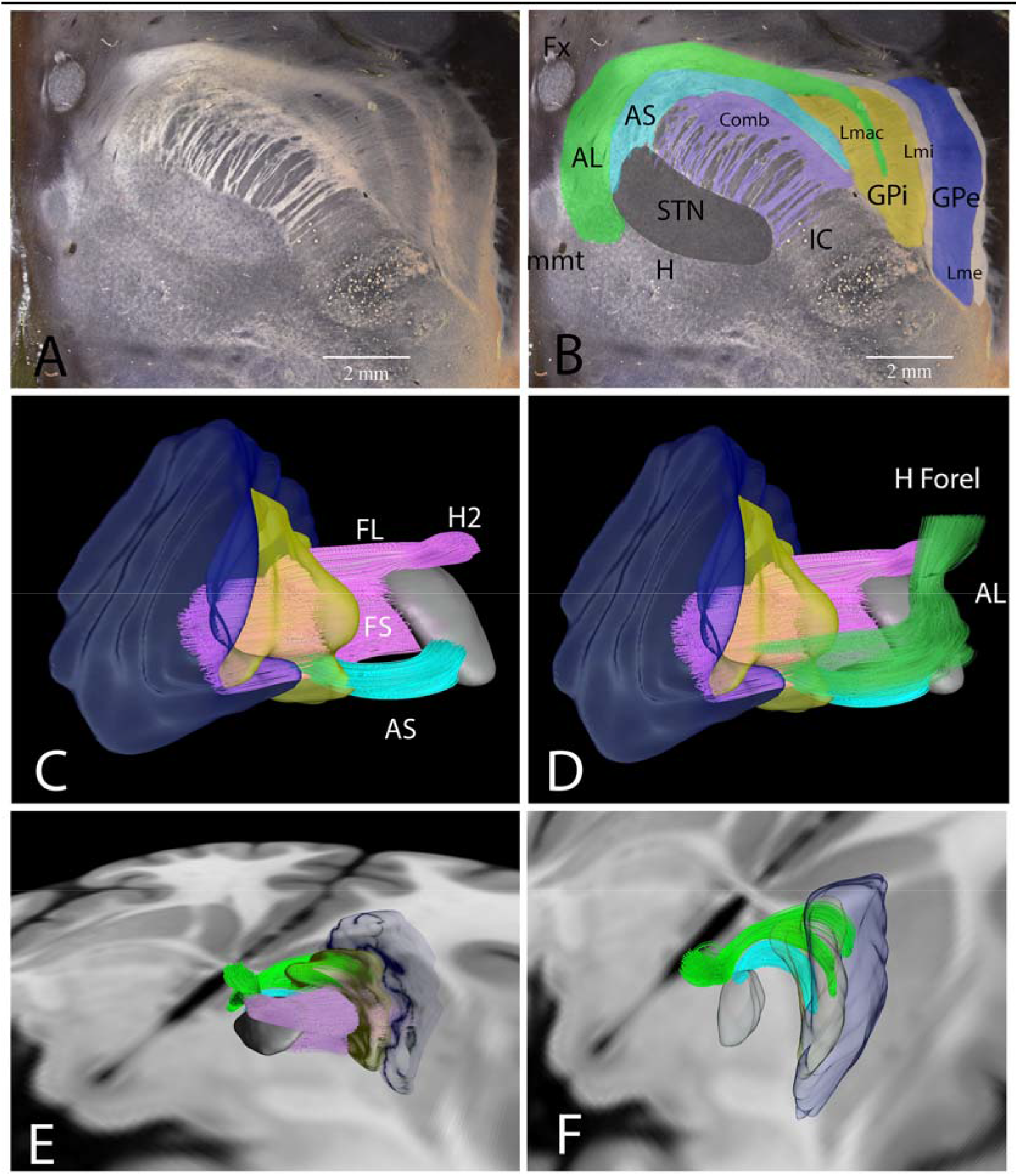
A) Horizontal 430 µm gallocyanin-stained section through the human right subthalamic region in darkfield illumination. B) Same histological section, labeled. The 3D reconstructions (C-E) follow the same color coding of the 2D slices. AL: ansa lenticularis (green); AS: Ansa subthalamica (light blue); H Forel: H field of Forel; Lmac: Lamina medullaris acessoria (green); Lmi: Lamina medullaris interna (white); Lme: lamina medullaris externa (white); Fx: fornix; mmt: mamilothalamic tract; Comb system (purple) composed by the fasciculus subthalamicus (FS) and fasciculus lenticularis (FL), that after crossing the dorsolateral border of the STN is referred as H2 field of Forel (H2); GPi: globus pallidus internus (yellow); GPe: globus pallidus externus (dark blue). C) Frontal 3D view of the right subthalamic region, showing the AS, the FS projecting to the dorsolateral STN from the GPe, the FL connecting the medial GPi to the H2 field of Forel. D) In the same view as C, the AL is added, and it is possible to see its course and confluence with the H2, to form the H Field of Forel. It is noticeable that in frontal planes of section the AS and AL can be confounded; E) 3D overview of the subthalamic region from a right-posterior pespective. F) The pallido-subthalamic and the Comb system were removed and it is possible to see the distinct paths of the AS and AL.

To demonstrate the bundles properties, we analyzed high-resolution dark field histological images in low and high power magnifications. In addition, we investigated *post-mortem* and *in vivo* ultra-high field (7T) imaging from 2 different imaging centers. Finally, in order to evaluate if the ansa subthalamica could be clearly identified in a routine clinical setting, we studied 13 pre-operative 3T MRI scans from idiopathic Parkinson’s disease (PD) patients.

## Methods

### Histological brain

The brain of a 56-year-old man with no history of neurological disorders was donated with written informed consent obtained from the family, according to the Declaration of Helsinki, as part of a research protocol approved by the Ethics board at the University of Sao Paulo. *In-situ* MRI scan was obtained before autopsy (T1: TR = 6.3 ms, TE = 2.9 ms, TI = 791 ms, 240 x 240, 1 x 1 x 1 mm voxels 3D).

The complete brain was fixed in formalin, embedded in celloidin and horizontally cut in 430 µm slices and stained using a modified gallocyanin technique^19^ as previously described^13^. We corrected tissue processing artifacts with multiple 2D and 3D registration procedures^20^ and finally registered the brain volume to a stereotactic common space (MNI ICBM152 2009b Nlin Asym), providing 3D coherent presentation of the neuroanatomic boundaries, segmented from the high-resolution dark-field microscopy images (resolution of 0.0250 x 0.0250 x 0.430 mm).

### High-field MRIs

High-field MRI was obtained in two different centers (University of Sao Paulo Medical School and the Athinoula A. Martinos Center for Biomedical Imaging, Massachusetts General Hospital). In the University of Sao Paulo, a 36-year-old man, with no previous diseases was scanned *in-vivo* in a 7T (Siemens Medical Solutions, Erlangen, Germany) device. Susceptibility-weighted Imaging (SWI) sequences with minimal intensity projection (mIP) reconstructions were obtained (0.64 mm isotropic voxels TR: 23ms, TE: 14 ms, FoV 174mm x 192mm, flip angle 12 degrees) and phase images show important contrast in the tegmental region (Fig. 2B).

**Fig. 2.**
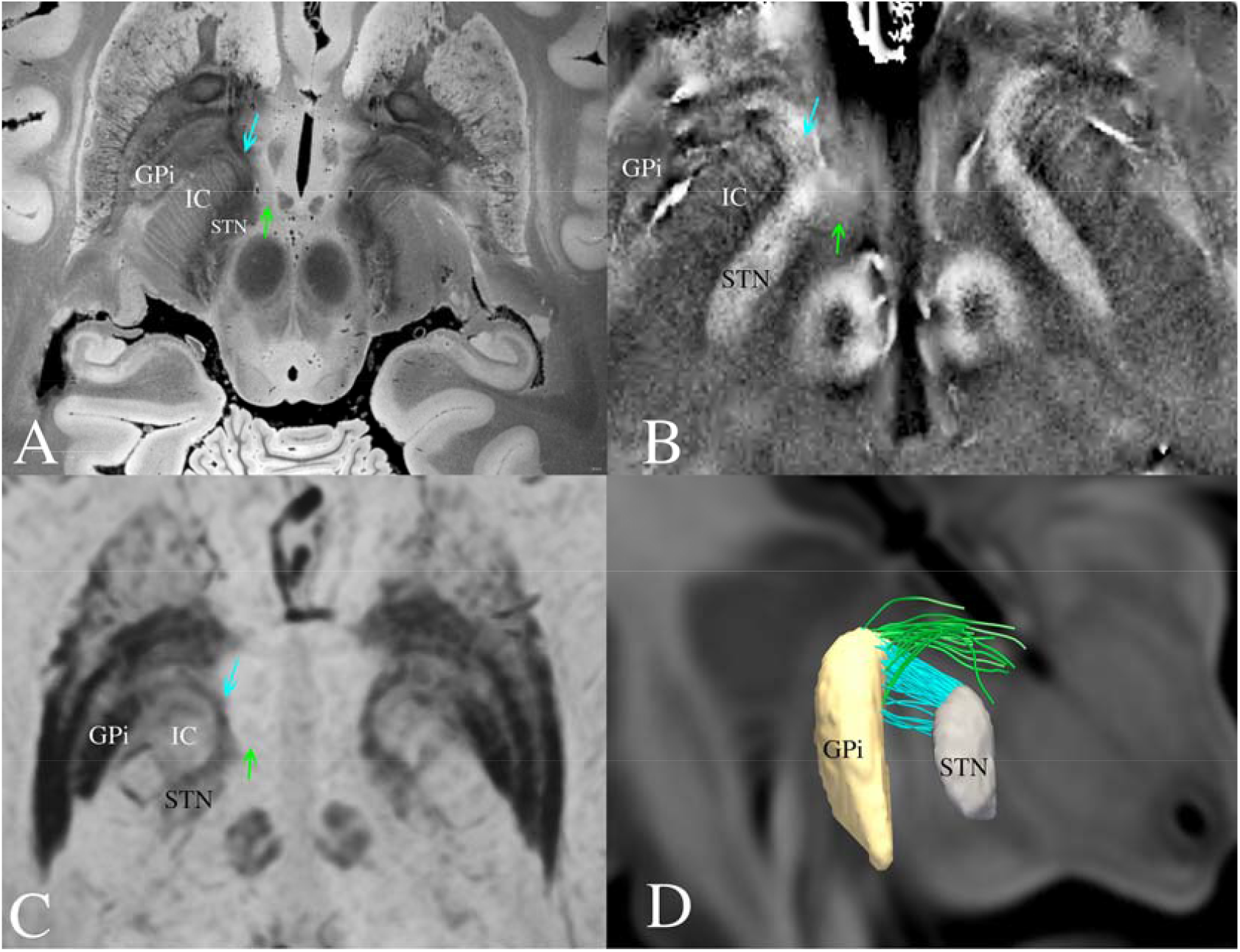
A) Post-mortem 7T MRI of a fixed human brain specimen. The axial slice is at the level of the rostral midbrain and diencephalon. It is possible to visualize the AS (blue arrow) curving around the Internal Capsule (IC) and the AL (green arrow) with different magnetic properties B) *In-vivo* 7T phase image of the same region C) Clincal miP-SWI image of a Parkinson’s disease patient, revealing the “horse shoe” shaped form of the AS, connecting the STN and GPi D) Connectome fibertracking showing different trajectories of AS (blue) and AL (green).

**Fig 3:**
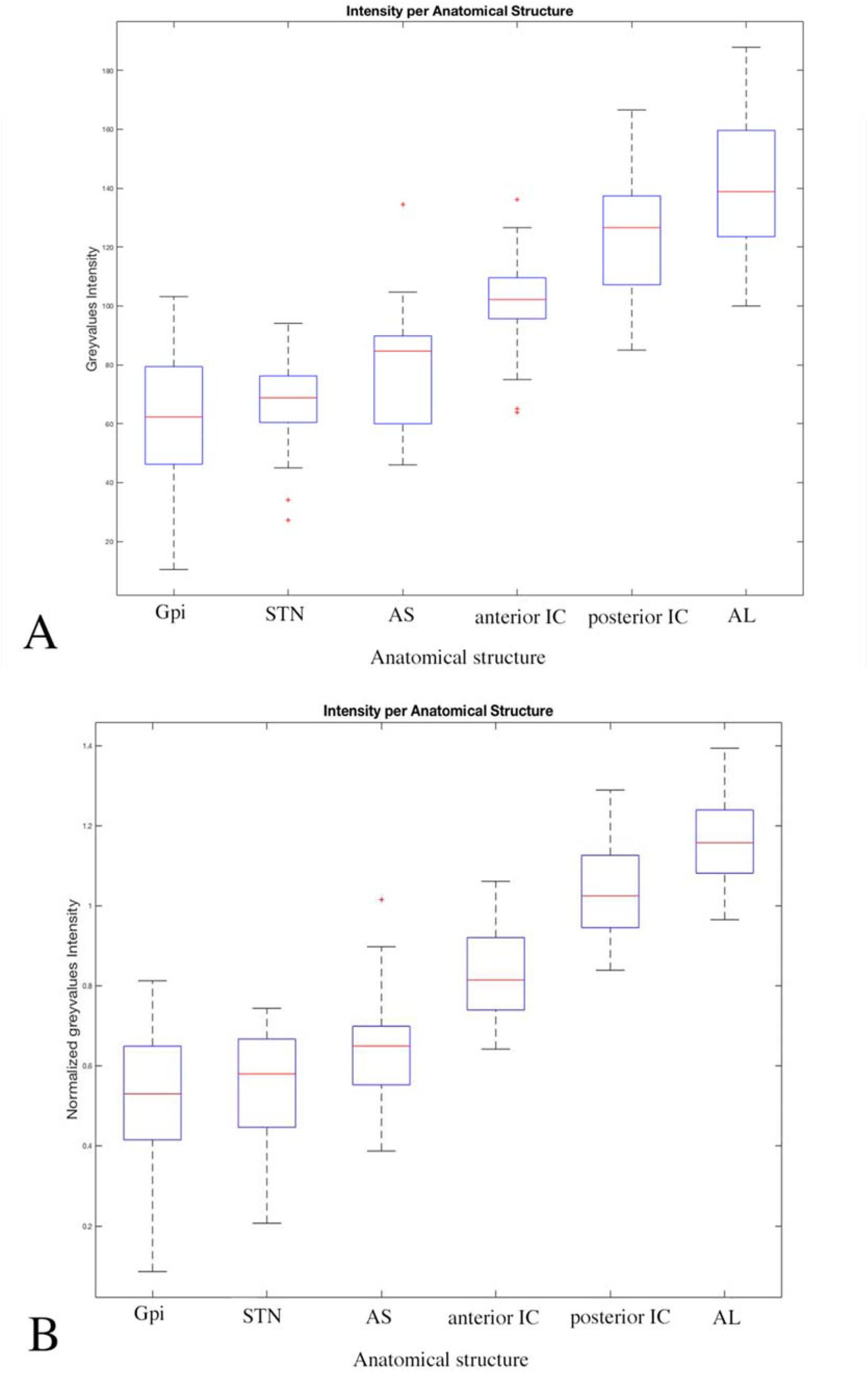
The grey values intensity for GPi, STN, ansa subthalamica (AST), anterior internal capsule (Ant IC), posterior internal capsule (Post IC) and Ansa lenticularis (AL) for 13 pairs of structures (n=26) is depicted. On each box, the central red mark indicates the median, and the bottom and top edges of the blue box indicate the 25th and 75th percentiles, respectively. The whiskers extend to the most extreme data points (not considered outliers), and the outliers are plotted individually using the red cross symbol. In A), GPi, STN and AS have the most hypointense signals, anterior IC has an intermediate signal and AL and the posterior IC have the most hyperintense signals in SWIw MRI. In part B), the normalized grey values removed outliers, but did not change any statistical diferences

At Massachusetts General Hospital (MGH), we scanned a normal brain specimen donated by a 60-year-old woman who died of non-neurological causes. Her neurological examination was normal during her last hospitalization prior to death, and the gross pathological analysis of her brain at autopsy was normal. The brain was fixed in 10% formalin 24 hours after death. The autopsy and *ex vivo* MRI analysis was performed with written informed consent provided by a surrogate decision-maker, in accordance with a research protocol approved by the Institutional Review Board at MGH.

We scanned the fixed whole brain specimen three months after the patient’s death on a 7T Siemens Magnetom MRI scanner (Siemens Medical Solutions, Erlangen, Germany) using a custom-built head coil. Details regarding the *ex vivo* scanning protocol have been previously published^21^. Briefly, we acquired a multi-echo FLASH (MEF) sequence^22^ at 200 µm spatial resolution. Total scan time on the 7T MRI scanner was 18 hours and 31 minutes. MEF data undergo a series of processing steps, including creation of parameter maps (e.g. proton density, flash20 and T1 and T2* relaxation times), which were used to synthesize images using the forward models of FLASH image formation to obtain high SNR data. For the present study, we analyzed the synthesized FLASH images using a 20 degree flip angle, based on visual inspection that revealed optimal delineation of the pallidofugal pathways.

### Clinical MRIs

Clinical anonymized MR images for pre-operative scans for DBS surgery in Parkinson’s disease patients were retrospectively analyzed. Thirteen patients were studied, 5 women and 8 men (age range: 38-75 years, mean age: 63 years, SD: 8,72 years). A T1 3D MPRAGE (0.90 mm isotropic voxels, TR: 2000 ms, TE: 2.4 ms TI: 925 ms, Flip angle: 8 degrees) and SW images (0.7 mm isotropic voxels, TR: 48 ms, TE: 30 ms, Flip angle 18 degrees) with mIP reconstructions were analyzed in the present study. Images were obtained on a 3T Siemens MRI scanner at Hospital Beneficencia Portuguesa de Sao Paulo (Fig. 2C).

### Three-dimensional histological reconstructions

The 3D reconstructions of the structures shown in Fig. 1C-F, were performed after manual segmentation on high-resolution histological dark-field images (Fig. 1A-B). The transformations generated by previous 2D, 3D and MNI registrations were then applied to the segmented masks. Nuclei were shown as solid structures and the fiber tracts were submitted to a technique called Histological Mesh Tractography (HMT). Briefly, inside 3D meshes that were segmented for each fiber tract, streamlines are generated using a finite volume method based simulation of waterflow in the computational fluid dynamics, using the software OpenFOAM® (https://www.openfoam.com/) and exported for visualization in the same stereotactic space as the original meshes.

### Clinical MRI analysis

Our primary goal was to verify if the ansa lenticularis and the ansa subthalamica had also different magnetic properties in SWI images due to differential iron content within each structure. To achieve that goal, the MRIs of the 13 patients were normalized to MNI ICBM152 2009b Nlin Asym space using Lead DBS software ^23^ and a modern three-stage non-linear method (Advanced Normalization Tools)^24^. We measured signal intensities in the histologically defined coordinates for the STN, GPi (already known to be rich in iron content^25^ and decreased signal in SW images), ansa subthalamica, ansa lenticularis, the anterior part of internal capsule (rich in perforating fibers) and the posterior part of internal capsule. The signal intensities were automatically normalized in SPM12 (https://www.fil.ion.ucl.ac.uk/spm/software/spm12/). To further normalize intensities, these were scaled using the anterior commissure as a reference point. The z-coordinate (−6.5 mm) was chosen to be the most representative axial slice of the ansa subthalamica.

The statistical analysis was performed for 13 MRI scans (n=26) with a one-way ANOVA test computed in the statistical JASP software (https://jasp-stats.org/). Post hoc Tukey’s test was performed and Cohen’s *d* factor was also calculated to verify respectively statistical difference among groups of structures and effect sizes of those differences (Table 1).

**Table:** Magnitude of the difference of the MRI normalized signal intensity values for the analyzed structures computed by Cohen’s *d* coefficient. The values are expressed in standard deviations. The smaller differences are seen between STN, GPi and AS (0,31-0,86), who share the most hypointense signals and between the AL and posterior IC (1,12), the most hyperintense structures. The anterior IC is in an intermediate signal zone. ANOVA *P-values* with 95% confidence interval of the difference between averages of MRI signal SWI intensity for each pair of structures. There was no evidence of difference for the average intensity of GPi and STN (p > 0.05), but for the comparisons of all the other analyzed structures, *p-values* were smaller than 0.05, suggesting a statistical difference of the mean MRI intensity signal.

| Magnitude of difference in MRI signal intensity and p-values with 95% of confidence interval |  |  |  |
| --- | --- | --- | --- |
|  |  | Cohen's d | p-values |
| **AL** | AS | 3.90 | < 0.001 |
|  | Anterior IC | 2.99 | < 0.001 |
|  | GPi | 4.30 | < 0.001 |
|  | Posterior IC | 1.12 | 0.013 |
|  | STN | 4.76 | < 0.001 |
| **AS** | Anterior IC | -1.36 | < 0.001 |
|  | GPi | 0.85 | 0.004 |
|  | Posterior IC | -2.92 | < 0.001 |
|  | STN | 0.64 | 0.162 |
| **Anterior IC** | GPi | 2.11 | < 0.001 |
|  | Posterior IC | -1.84 | < 0.001 |
|  | STN | 2.13 | < 0.001 |
| **GPi** | Posterior IC | -3.45 | < 0.001 |
|  | STN | -0.31 | **0.766** |
| **Posterior IC** | STN | 3.73 | < 0.001 |

## Results

Figures 1A and B depict the ansa subthalamica as a conspicuous bundle of fibers coursing between the antero-medial pole of the STN and the ventro-medial portions of the GPi, at levels below the anterior commissure. The ansa lenticularis was composed of white matter fibers derived from the accessory medullary lamina and from the internal medullary lamina, whereas the pallidal portion of this bundle arches adjacent to the medial to border of the GPi, and lateral to the Comb system. These fibers form a separate bundle connecting the anteromedial pole of the STN with the medial portions of the pallidum. A 3D visualization of the intricate relationship of these structures is shown in Fig. 1C-F.

These anatomical aspects were verified in the *post-mortem* and *in-vivo* high-field MRI (figure 2). Three-dimensional fibertracking based on a high-definition normative connectome from human data confirmed findings noninvasively (figure 2D).

The mean intensity values in SW images for the measured regions were lower for the GPi (61.08 ± 23.85), STN (66.47 ± 15.51) and ansa subthalamica (78.71 ± 21.03), intermediate for the anterior internal capsule (99.85 ± 16.76) and higher for the posterior internal capsule (125.30 ± 21.13) and ansa lenticularis (140.70 ± 22.67 (Graphic 1A). Tukey’s test p value was bigger than 0.05 for STN x ansa subthalamica and STN x GPi comparisons, meaning the 3 structures belong to one single group regarding grey values intensities in SW images. All the other comparisons showed there was a significant statistical difference (p<0.05) between groups (Table).

Effect sizes in differences of image intensities expressed by Cohen’s *d* factors showed that GPi, STN and ansa subthalamica have smaller differences among them (−0.31 to 0.86 SD), while the anterior IC is in an intermediate position between the GPi/STN/ansa subthalamica group and the posterior IC/ansa lenticularis group. Posterior IC and ansa lenticularis have smaller differences between them (1.12 SD), but larger differences to GPi, STN and ansa subthalamica (3.90 to 4.76 SD) (Table 1). The analyzed regions could be divided in 3 groups: hypointense (GPi, STN, and ansa subthalamica), isointense (anterior IC) and hyperintense (posterior IC and ansa lenticularis) in susceptibility weighted images.

## Discussion

Since Auguste Forel’s original descriptions in 1877^26^, much of the classical literature on the tegmental area has focused on confined assessment of coronal planes of sections in humans. On a coronal plane, the ansa subthalamica and ansa lenticularis can be mistaken from one another (Fig. 1D). Riley characterized the AS as a subdivision of the AL under the term *fibrae pallido-subthalamicae*^18^. The ansa subthalamica, however, differs anatomically from the ansa lenticularis on MRI intensity, histological properties and trajectory (Fig. 1A-F). While the ansa lenticularis traverses from the GPi to the thalamus, the ansa subthalamica runs from the GPi to the STN. We thus argue that the two are separate and distinguishable pathways and the ansa subthalamica is not part of the ansa lenticularis.

The susceptibility weighted Imaging (SWI) is particularly sensitive to paramagnetic and diamagnetic compounds such as deoxyhemoglobin, ferritin, bone minerals and calcifications, causing drop of signal. Pallidofugal and striatonigral pathways have been visualized *in vivo* using SWI contrasts and the tubular structures in the vicinity of the STN and substantia nigra proved to represent fiber tracts rather than veins in previous studies^27^. The hypointensity in the basal ganglia regions in SWI is probably due to the presence of iron^25^. Minimum intensity projection (mIP) images augment this effect for smaller regions. The signal is similar to STN and GPi within the ansa subthalamica. Instead, the ansa lenticularis was hyperintense (likely due to poor iron content), similar to the posterior portions of the IC. The intermediate signal of the anterior IC might be explained by the FS projections that traverse through the internal capsule and reach the STN (Edinger’s comb system). Here, iron-rich projections cross the iron-poor IC, generating an intermediate signal. The hypointense signal from the ansa subthalamica also has a ventral projection to the substantia nigra. These projections from GPi to the SNr first follow the course of the ansa subthalamica, but then extend ventrally. Basal ganglia connectivity studies have confirmed the presence of the connections between STN and GPi as well as GPi and SNr using high-field MRI in humans^28,29^, but no detailed anatomical description was provided.

The ansa subthalamica connects the anterior and ventral parts (below the anterior commissure level) of the pallidum, to the anterioventral part of the STN ^1^-both of which have been attributed to predominantly limbic processing. This suggests that the ansa subthalamica may primarily convey limbic information. Instead, motor connections of the STN to GP are likely implemented in the SF, on the dorsolateral aspects of the STN^1^. Given the widely described cross-talk between motor-, associative and limbic loops in the basal ganglia and on level of the cortex, the ansa subthalamica may still play a crucial part within the fronto-striato-subthallamo-pallidal network engaged in goal directed behaviors and habitual inhibition^30^. These implications, which presently remain speculative, have potential translational relevance in the field of deep brain stimulation to the subthalamic region in motor and psychiatric disorders.

In summary, we provide a comprehensive description of the ansa subthalamica as a distinct fiber bundle from the ansa lenticularis, lenticular fasciculus and subthalamic fasciculus in the human pallido-subthalamic circuitry. The ansa subthalamica connects the antero-medial STN to the ventral aspects of the GPi and is recognizable not only in histological preparations but also on high-field MRI, connectome tractography, and in clinical MRI scans of PD patients in iron-sensitive sequences. It may represent the anatomical pathway that connects limbic regions of the STN and pallidum, and should be investigated as a possible substrate for the effects of stereotactic surgery, particularly in psychiatric disorders.

## Data Availability

Data are available upon reasonable request

## Abbreviations

AL: ansa lenticularis
AS: ansa subthalamica
DBS: Deep brain stimulation
GPe: globus pallidus externus
GPi: globus pallidus internus
IC: Internal capsule
SF: subthalamic fasciculus
STN: subthalamic nucleus
LF: lenticular fasciculus

## Acknowledgement

We are grateful to the volunteers who participated and Family members who donated the brain for this study. We would also like to thank all the members of the São Paulo-Würzburg collaborative Project. This includes the participants of the Brain Bank of the Brazilian Aging Brain Study Group, Mrs. E. Broschk and Mrs A. Bahrke from the Morphological research Unit of the University of Würzburg.

## Authors’ Roles

1) Research project: A. Conception, B. Organization, C. Execution; 2) Statistical Analysis: A. Design, B. Execution, C. Review and Critique; 3) Manuscript: A. Writing of the first draft, B. Review and Critique.

Alho EJL: 1) A,B,C; 2)A,B; 3) A

Alho ATDL: 1)B,C; 3)B

Horn A: 1) C; 2)C 3)B

Martin MGM: 1)C 2)C 3)B

Edlow BL: 1)C 2)C 3)B

Fischl B: 1)C 2)C 3)B

Nagy J: 1)C 2)C 3)B

Fonoff ET: 1)C 2)C 3)B

Hamani C: 1)C 2)C 3)B

Heinsen H: 1)A,C 2)C 3)B

